# Acceptability and efficacy of vaginal self-sampling for genital infection and bacterial vaginosis: A large, cross-sectional, non-inferiority trial

**DOI:** 10.1101/2021.01.05.21249269

**Authors:** Camus Claire, Penaranda Guillaume, Khiri Hacène, Camiade Sabine, Molet Lucie, Lebsir Melissa, Plauzolles Anne, Chiche Laurent, Blanc Bernard, Quarello Edwin, Halfon Philippe

## Abstract

**Objective:** Screening for genital infection (GI), bacterial vaginosis (BV), sexually transmitted infection (STI) and asymptomatic carriage of group B streptococcus (GBS) in pregnant women is a common reason for medical appointments. Objectives were first to determine the non-inferiority of vaginal self-sampling compared with vaginal/cervical classical sampling to screen for GIs, bacterial vaginosis (BV), STIs, and GBS asymptomatic carriage in pregnant women; second to determine the feasibility of vaginal self-sampling.

**Methods:** Vaginal self-sampling (VSS) and vaginal/cervical classical sampling (VCS) of 1027 women were collected by health care professionals and simultaneously carried out on each patient. Bacterial infection, yeast infection, Chlamydia trachomatis, Neisseria gonorrhoea, Mycoplasma genitalium, Trichomonas vaginalis and Herpes simplex virus types were systematically screened in both paired VSS and VCS samples.

**Results:** Statistical tests supported the non-inferiority of VSS compared with VCS. Agreements between VCS and VSS remained high regardless of the type of studied infection. VSS had successful diagnostic performances, especially for Predictive negative value (PNV) (over 90%) for all studied infections. Most participants (84%) recommended the use of VSS.

**Conclusions:** This study remains the most exhaustive in screening for GI, BV, STI agents and asymptomatic GBS carriage. Given its efficacy and acceptability, VSS seems to be a viable alternative to classic physician sampling among women in the general population. This study provides evidence that vaginal self-sampling can be used as a universal specimen for detection of lower genital tract infections in women.

**Study Identification number:** ID-RCB 2014-A01250-4

## INTRODUCTION

Screening for genital infection (GI), bacterial vaginosis (BV), sexually transmitted infection (STI) and asymptomatic carriage of group B streptococcus (GBS) in pregnant women is a common reason for medical appointments. GI including bacterial vaginosis and yeast colonization are the most common lower genital tract disorders among women of reproductive age ^1,2^. STIs are a major public health concern, with Chlamydia trachomatis (CT), Neisseria gonorrhoeae (GC) and genital herpes (Human simplex virus, HSV) the most prevalent STI and Mycoplasma genitalium (MG) an emerging sexually transmitted pathogen ^3,4^. The diagnosis and control of GIs, BVs, and STIs are important issues, given their long-term consequences for fertility and overall well-being of affected women ^1,4^. In the same way, intrapartum screening for GBS colonisation is recommended in France at 35 to 37 weeks of gestation to initiate antibiotic prophylaxis and prevent early-onset infection in neonates ^5,6^.

In current medical practice, vaginal and/or cervical sampling is indicated in cases of suspicion of (symptomatic) or screening for (asymptomatic) GIs, STIs or cases of asymptomatic carriage in pregnant women in the eighth month of pregnancy. Conventional testing is performed by using vaginal and/or cervical classic sampling (VCS). This procedure requires a pelvic examination, speculum use and vaginal or endocervix sampling by health care professionals such as gynaecologists, general practitioners, medical biologists and midwives. Concerns related to cultural and religious norms, as well as economic issues, are often cited as barriers to screenings performed with this method ^7–10^. Moreover, specimen collection is often complicated by in-clinic difficulties, such as delayed waiting time to obtain a medical appointment and the lack of practitioners ^11^. Vaginal self-sampling (VSS) has progressively emerged as an alternative to VCS, primarily for human papillomavirus (HPV)–associated cervical cancer screening. Previous studies have already been conducted by several teams, including us, and shown the efficacy and acceptability of VSS in this indication ^12–16^. More recently, VSS was used for STI agent screening, and was recommended by participants of workshop conducted by the National Institutes of Health (NIH) for CT and NG screening ^8,17^. Indeed, the ease and acceptability of vaginal self-sampling could facilitate follow-ups and potentially help in the prevention of gynaecological disorders.

By performing a large, randomised, cross-sectional study, our objectives were to:

1. Determine the non-inferiority of VSS compared with VCS for the detection of GI, BV, STI infections and GBS asymptomatic carriage in pregnant women.
2. Determine the feasibility of self-collection by women requiring vaginal and/or cervical sampling for their gynaecological monitoring.

## MATERIALS & METHODS

### Sample size calculations

Sample size was calculated to demonstrate non-inferiority of VSS compared with VCS for the detection of each infections separately: GI, BV, STI, and GBS asymptomatic carriage in pregnant women. The worst-case scenario was calculated for CT infection: considering a prevalence of 4.4% [95% Confidence Interval (95% CI) 2.1-8.0%] for VCS (P_VCS_) (results observed in routine laboratory analysis from 225 patients in 2014), and a non-inferiority margin (M) of 2.3% (defined as the difference between P_VCS_ and 95% CI lower bound of P_VCS_). Under these conditions, if there is no difference between P_VCS_ and P_VSS_, then at least 984 patients were required to be sure that the upper limit of a one-sided 95% CI excluded a difference in favour of VCS of more than 2.3% (alpha level of 5%, and statistical power of 80%) ^18^.

### Study population

From October 2015 to March 2018, 1027 participants, including 224 pregnant women (21.8%), were recruited from 11 clinical centres in Marseille, France. Recruitment was conducted by 26 clinical practitioners including gynaecologists and midwives. Women were eligible to participate if they were 18–65 years of age and if they presented vaginal/cervical sampling indications (e.g., suspicion or screening for GI and/or STI and/or asymptomatic carriage of GBS, especially for pregnant women). At recruitment, clinical practitioners filled out a Clinical information form where sampling indications were mentioned. In addition, participants were asked to complete a questionnaire to collect their feedback toward self-sampling and classic sampling (Supplementary File 1).

Written informed consents were obtained from the participants at recruitment. The study was sponsored by the Hôpital Européen Marseille Clinical Research Department in collaboration with the Clinical Research and R&D Department of the Laboratoire Européen Alphabio. This biomedical clinical study was authorised by the French competent authority (ANSM, www.ansm.sante.fr) and received the agreement of an ethics committee (CPP Sud Méditerranée I). The French national identification number of the study is ID-RCB 2014-A01250-47.

### Sample collection

Participants were asked to provide two types of genital samples:

1. Self-collected vaginal swabs (named “vaginal self-sampling” or VSS).
2. Vaginal/cervical swabs collected by a medical practitioner during a pelvic examination (named “vaginal classic sampling” or VCS).

Randomised sampling technique was performed as follows: Half of the women were asked to perform VSS before VCS, and the other half were asked to perform VSS after VCS. Sampling order was specified on the clinical information form. A VSS paired sampling (randomised order) and a VSS acceptability survey were prospectively collected.

Performing their VSS, participants were instructed to push the swab gently into their vagina, turn it two times while wiping the vaginal wall and slowly remove the swab. The swab was then inserted and released into the collection tube prior to capping with the lid. VCSs were collected following the same procedure under speculum examination.

Two swabs were collected for both VSS and VCS: Copan ESwab (Biomérieux, Marcy l’Etoile, France) and Cobas PCR media (Roche Diagnostics, Meylan, France). Microbiological analysis and HSV molecular biology analysis were performed using Copan ESwab, and TV, CT, NG and MG molecular biology analysis were performed using Cobas PCR media.

### Microbiological analysis

The VSS and VCS tubes were shipped at ambient temperature to Alphabio laboratories in Marseille for microbiological analysis and STI testing. Both VSS and VCS were treated within 24 h of sample collection. For each of the paired samples, routine microbiology analysis was performed : BV was defined by a Nugent score equal or superior to 7, and yeast colonization was defined by yeast growth on nutrient agar media^19^. In addition, a large panel of nucleic acid amplification tests (NAAT), targeting STI agents, were performed: CT, NG (Cobas, Roche Diagnostic), MG, TV (Roche Diagnostic, TIBMolBiol) and HSV types 1 and/or 2 (Cobas 4800 Roche Diagnostic). GI referred to bacterial and/or yeast infection, and STI infection referred to CT and/or NG and/or MG and/or TV and/or HSV.

### Statistical analysis

Data were reported using frequencies and proportions. Non-inferiority of VSS compared with VCS was assessed using z statistic for binomial proportions. The null hypothesis for the non-inferiority test is H0: P_VSS_ – P_VCS_ ≤-M versus the alternative H1: P_VSS_ – P_VCS_ >-M, where P_VSS_ stands for the prevalence observed for VSS, P_VCS_ stands for the prevalence observed for VCS and M stands for the non-inferiority margin defined as the 95% confidence interval [CI95] lower bound of P_VCS_. Exact Clopper–Pearson CI95s were reported. Overall agreement between VCS and VSS was reported, calculating the rate of concordant cases, kappa statistic, positive predictive value (PPV), negative predictive value (NPV), sensitivity (Se) and specificity (Sp) (all with their 95% CI)^20^; the McNemar test for matched paired data was computed to test the marginal homogeneity between VCS and VSS. Sampling randomisation sequences were compared using the Breslow-Day test (VCS before VSS vs. VSS before VCS) assuming equality of odds ratios (null hypothesis). Statistical tests were assessed using a significance criterion of α = 0.05. Statistical computations were performed using SAS v. 9.4 software (SAS Institute, Inc., Cary, NC).

## RESULTS

### Sampling indications

Mean age was 35.2 (standard deviation 11.5) years old. Indications of genital sampling comprised suspicion of GI in 380 participants, suspicion of STI in 62 participants, suspicion of or screening for GI plus STI in 419 participants, and GBS asymptomatic carriage in 166 pregnant women (Table 1).

**Table 1.**
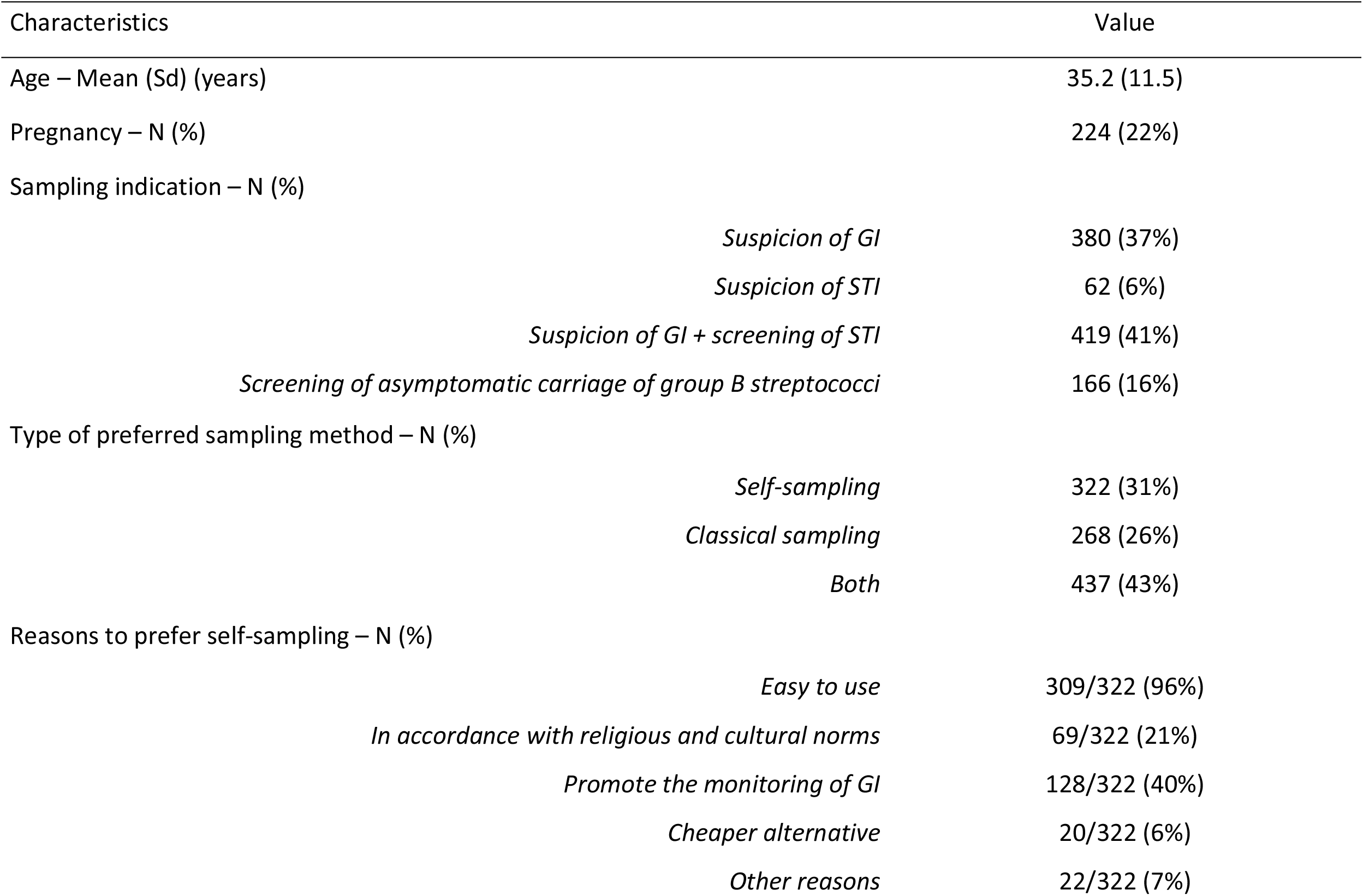
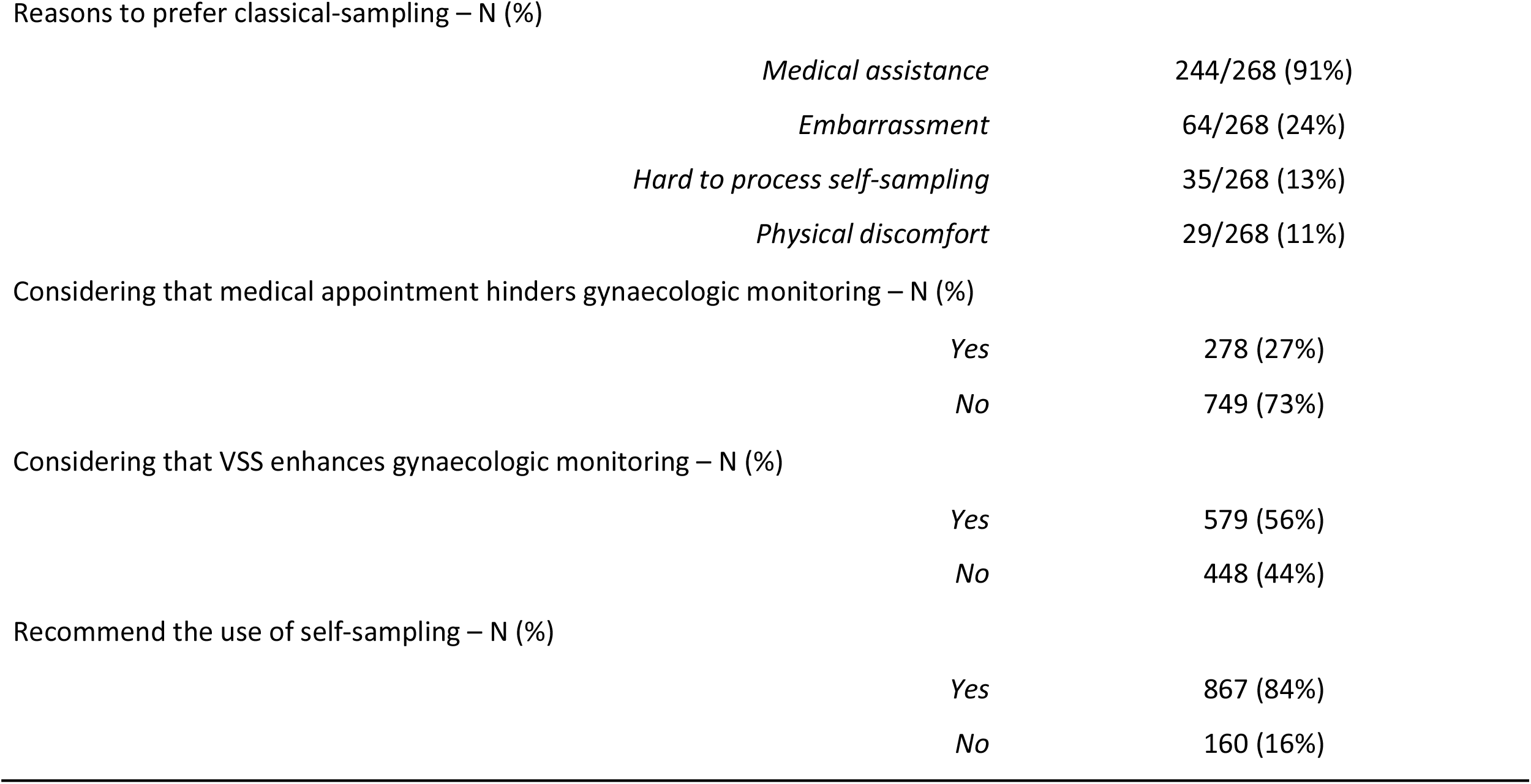
Characteristics of surveyed women

| Characteristics | Value |
| --- | --- |
| Age – Mean (Sd) (years) | 35.2 (11.5) |
| Pregnancy – N (%) | 224 (22%) |
| Sampling indication – N (%) |  |
| *Suspicion of GI* | 380 (37%) |
| *Suspicion of STI* | 62 (6%) |
| *Suspicion of GI + screening of STI* | 419 (41%) |
| *Screening of asymptomatic carriage of group B streptococci* | 166 (16%) |
| Type of preferred sampling method – N (%) |  |
| *Self-sampling* | 322 (31%) |
| *Classical sampling* | 268 (26%) |
| *Both* | 437 (43%) |
| Reasons to prefer self-sampling – N (%) |  |
| *Easy to use* | 309/322 (96%) |
| *In accordance with religious and cultural norms* | 69/322 (21%) |
| *Promote the monitoring of GI* | 128/322 (40%) |
| *Cheaper alternative* | 20/322 (6%) |
| *Other reasons* | 22/322 (7%) |
| Reasons to prefer classical-sampling – N (%) |  |
| *Medical assistance* | 244/268 (91%) |
| *Embarrassment* | 64/268 (24%) |
| *Hard to process self-sampling* | 35/268 (13%) |
| *Physical discomfort* | 29/268 (11%) |
| Considering that medical appointment hinders gynaecologic monitoring – N (%) |  |
| *Yes* | 278 (27%) |
| *No* | 749 (73%) |
| Considering that VSS enhances gynaecologic monitoring – N (%) |  |
| *Yes* | 579 (56%) |
| *No* | 448 (44%) |
| Recommend the use of self-sampling – N (%) |  |
| *Yes* | 867 (84%) |
| *No* | 160 (16%) |

### Detected infections using vaginal self-sampling and concordance with vaginal classical sampling

Concordance between VSS and VCS was assessed among 1026 samples as one VCS failed for molecular biology analysis. The prevalence of overall STI was 8.5% using VSS, and 8.1% using VCS (Table 2). Prevalence of specific infections were respectively 1.9% (VSS) versus 1.8 % (VCS) for TV, 3.3% (VSS) versus 3.2% (VCS) for CT, 1.0% (VSS) versus 0.9% (VCS) for NG, 1.4% (VSS) versus 1.4% (VCS) for MG and 1.9% (VSS) versus 1.7% (VCS) for HSV showing non-inferiority of VSS compared with VCS; and kappa values computed for STI were all superior or equal to 0.95, which indicates excellent agreement between self-collected and physician-collected samples. NcNemar test for marginal homogeneity was not significant, showing that marginal frequencies are not statistically different.

**Table 2.** Detection rates of GI, STI infection and GBS asymptomatic carriage, and non-inferiority test Vaginosis detection rates were determined among patients with bacterial vaginosis and/or yeast colonization. STI detection rates were determined among patients with TV, and/or CT, and/or NG, and/or MG, and/or HSV infection.

| Sample type | Vaginal classic-sampling |  | Vaginal self-sampling |  | P-value (for non-inferiority of vaginal self-sampling) |
| --- | --- | --- | --- | --- | --- |
|  | Positive<br>N | % (95% CI) | Positive<br>N | % (95% CI) |  |
| GI | 392 | 38.1 (35.2-41.1) | 408 | 39.7 (36.7-42.7) | 0.0016 |
| Bacterial vaginosis | 105 | 10.7 (8.8-12.7) | 104 | 10.6 (8.7-12.5) | 0.0348 |
| Yeast colonization | 304 | 29.6 (26.8-32.4) | 322 | 31.3 (28.5-34.2) | 0.0009 |
| STI | 83 | 8.1 (6.4-9.7) | 87 | 8.5 (6.8-10.2) | 0.0087 |
| Trichomonas vaginalis | 18 | 1.8 (1.0-2.6) | 19 | 1.9 (1.0-2.7) | 0.0217 |
| Chlamydia trachomatis | 33 | 3.2 (2.1-4.3) | 34 | 3.3 (2.2-4.4) | 0.0354 |
| Neisseria gonorrhoeae | 9 | 0.9 (0.3-1.4) | 10 | 1.0 (0.4-1.6) | 0.0140 |
| Mycoplasma genitalium | 14 | 1.4 (0.7-2.1) | 14 | 1.4 (0.7-2.1) | 0.0336 |
| Herpes simplex virus | 17 | 1.7 (0.9-2.4) | 19 | 1.9 (1.0-2.7) | 0.0120 |
| Group B streptococcus | 118 | 11.5 (9.5-13.2) | 138 | 13.4 (11.3-15.5) | 0.0001 |
| Group B streptococcus in pregnant women | 18 | 8.0 (4.5-11.6) | 20 | 8.9 (5.2-12.7) | 0.0010 |

Prevalence of GIs including BV and yeast colonization were respectively 39.7% (VSS) versus 38.1% (VCS), 10.6% (VSS) versus 10.7% (VCS) and 31.3% (VSS) versus 29.6% (VCS); showing no inferiority of VSS to VCS (Table 2). Prevalence of GBS asymptomatic carriage was 8.9% (VSS) versus 8% (VCS) in pregnant women. Agreements between VCS and VSS remained high ranging from 90.3% for GI up to 98.3% for GBS asymptomatic carriage in pregnant women, and kappa statistics showed substantial agreement between VCS and VSS (ranging from 0.77 for BV to 0.89 for SGB asymptomatic carriage in pregnant women). McNemar test for marginal homogeneity was not significant except for yeast colonization and GBS (p = 0.0442 and p = 0.0004, respectively).

With VCS as the standard reference, VSS had successful diagnostic performances, especially the NPVs over 90% for all studied infections (table 3).

**Table 3.**
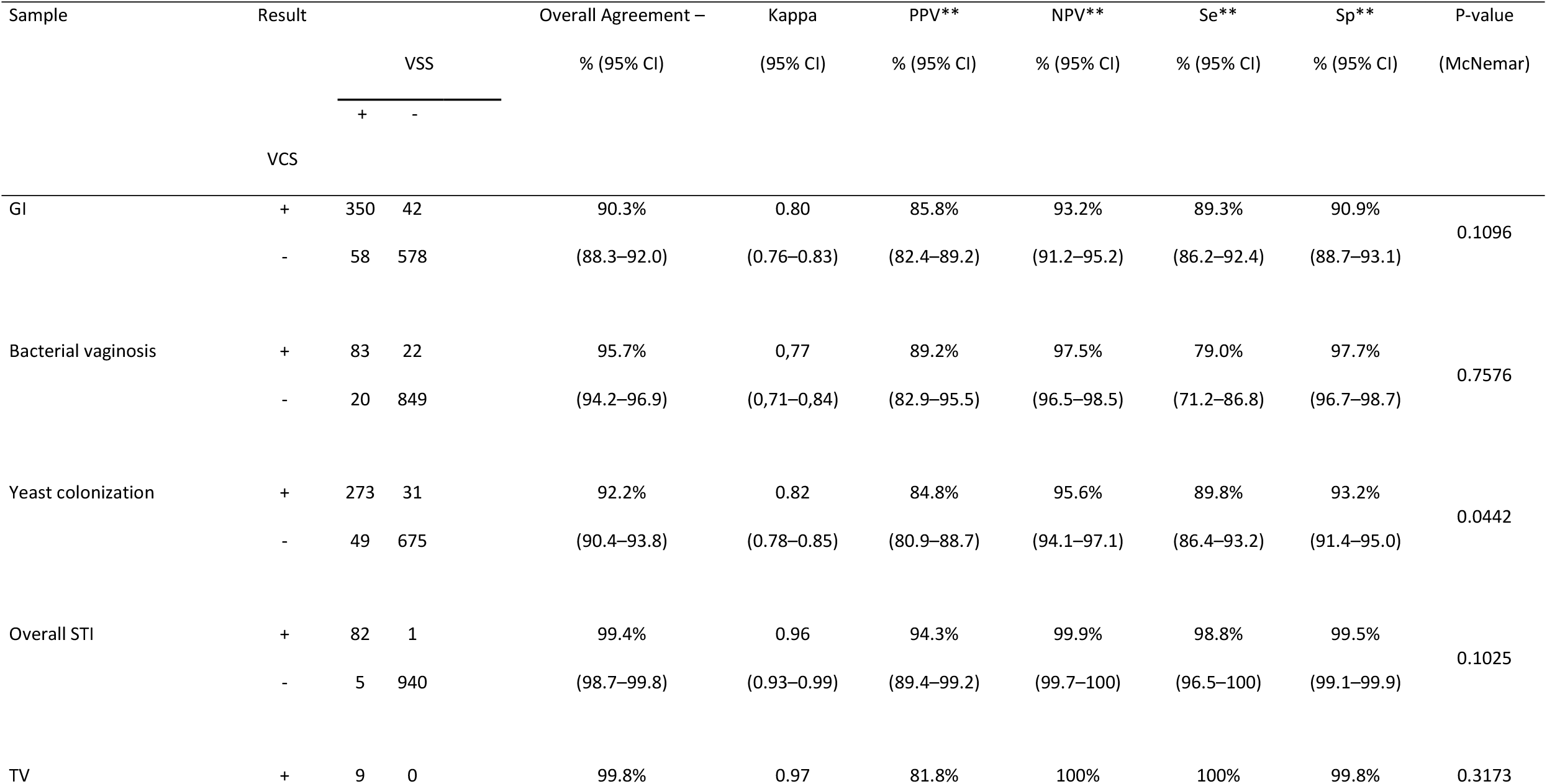
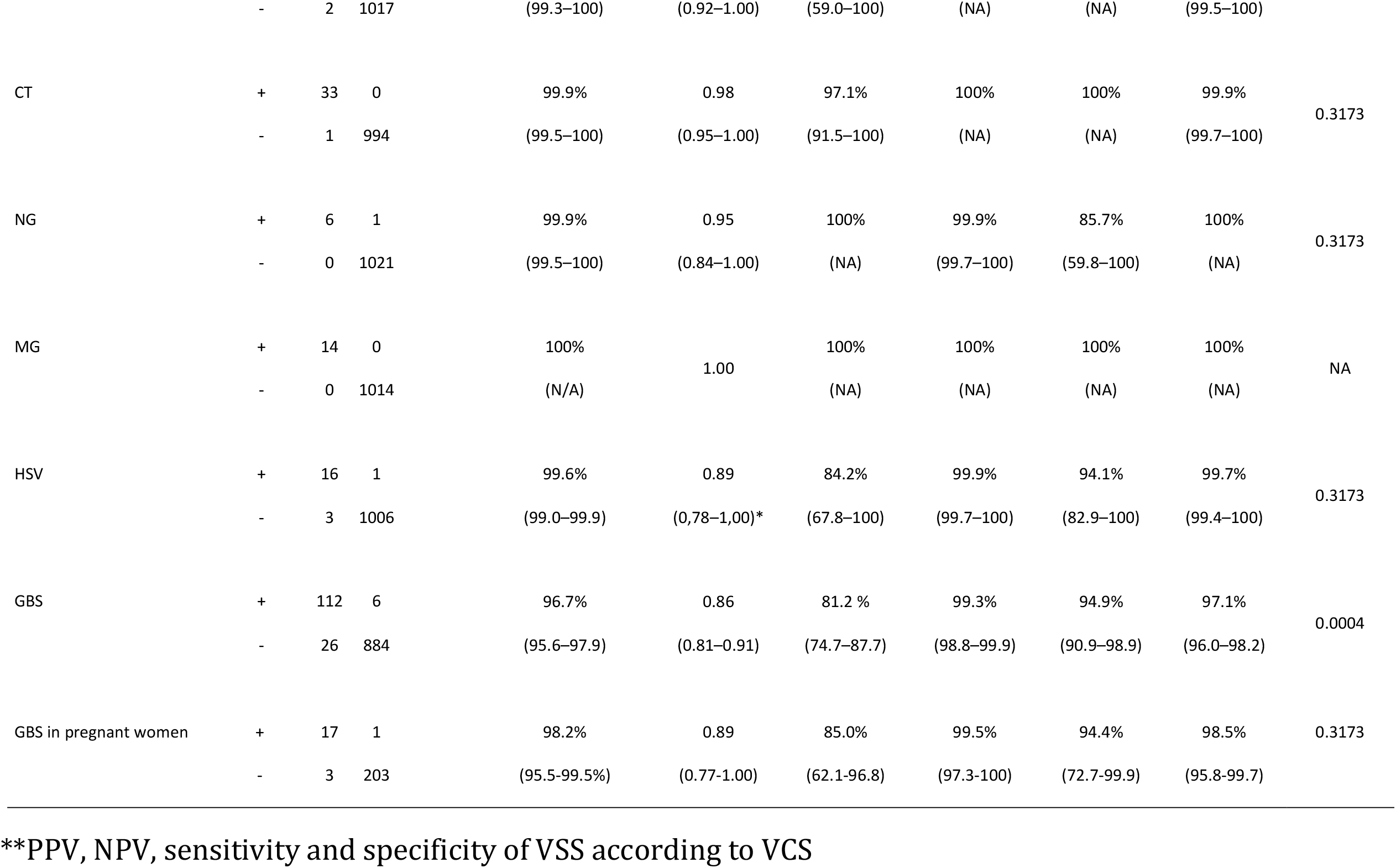
Agreement between VSS and VCS for the detection of GI agents *Agreement between VSS and VCS were determined among 1026 patients (one VCS failed and was excluded from statistical analysis).

### Attitude toward self-sampling and clinical collection

Most women expressed a preference for VSS compared with VCS: 322 (31%) vs. 268 (26%) respectively, p = 0.045 (437 (43%) women did not express a preference between VSS and VCS) (Table 1). The primary reason women gave for preferring VSS was its ease of use: 96% (309/322). Other reasons to prefer VSS were as follows: 40% (138/322) of them estimated that VSS facilitated monitoring of GI, 21% considered VCS in accordance with their cultural norms and 6% considered VSS a cheaper alternative to VCS. The main reason women preferred VCS was because they preferred to be sampled by a practitioner: 91% (244/268). Other reasons to prefer VSS were as follows: 24% reported embarrassment, 11% reported physical discomfort performing VSS and 13% reported that it was difficult to perform VSS. Of the 1027 surveyed participants, 84% (867) would recommend the use of VSS, and 56% (579) asserted that VSS instead of VCS would encourage them to be monitored more regularly. In contrast, only 27% (278) of participants considered the need to have a medical appointment to perform vaginal sampling as an obstacle to gynaecological monitoring.

## Discussion

### Principal Findings

The principal findings and interpretations of the study are as follows : 1) VSS has proved its efficacy for all tested infections, including GI, BV, STI and GBS asymptomatic carriage; 2) Statistical tests supported the non-inferiority of VSS compared with VCS; 3) Agreements between VCS and VSS remained high regardless of the type of studied infection; 4) Acceptability of VSS was satisfactory among women from the general population in a global setting; 5) VSS should be proposed to women from the general population in a clinical setting; 6) The use of VSS should enhance gynaecological monitoring. This provide evidence that VSS would be an effective alternative to VCS.

### Efficacy of VSS in screening infectious agents

Efficacy of VSS was evaluated for a large panel of infectious agents. Vaginal self-sampling was comparable with vaginal classic sampling for CT, NG, MG and TV detection by NAAT, consistent with findings in other cohorts ^10,17,21–25^. CT, NG, MG and TV detection rates were lower than those previously reported in under-screened low-income women^4,26^ and might be influenced by recruitment. In fact, women participating in this study were recruited by clinicians during their course of care and STIs were suspected in only 6% of participants, as has been mentioned before. HSV detection rates were also very low, consistent with intermittent viral shedding in the mucosa of infected populations ^27^. The close agreement (99.6%; [99-99.9]) between VSS and VCS detection rates encourages the use of VSS to detect any HSV active infection.

GI and GBS asymptomatic carriage were screened by microbiological analysis. The microbiological analysis implies another issue compared to NAAT: it should be rapidly performed to maintain bacterial viability and avoid false negative results. In this study, both VSS and VCS were processed within a maximum of 24 hours, thus ensuring good analytical conditions. Prevalence of bacterial vaginosis and yeast colonization were similar to previously reported ^28–36^, and self-collection was comparable to clinician-based collection. These results showed that self-collected vaginal swabs are an effective method for the diagnosis of both bacterial vaginosis and yeast infection. Some papers support the existing knowledge regarding GBS screening by vaginal self-sampling. Several authors showed the non-inferiority of VSS compared to VCS ^1,37–40^. One study reported contradictory results, demonstrating that self-collection was less sensitive to detect GBS carriage ^41^. In this study good kappa values were reported, eg. 0.86 and 0.89 in pregnant women, establishing the good reliability of self-collected vaginal swabs to screen GBS asymptomatic carriage. In addition, sensitivity was 94.4% in pregnant women, allowing the use of VSS in this indication. Given its excellent diagnostic performance, especially for high NPV, health care practitioners can be confident in the use of VSS to detect any symptomatic or asymptomatic infection.

### Overall acceptability of VSS among women

The principal indication was suspicion of GI (bacterial and yeast infection), which represented 77.8% (800) of the study’s participants. GI screening was combined with STI screening in 420 women (52%). In fact, GI, particularly BV, have been found to be associated with sexually transmitted agents such as NG, CT and MV ^1,42^. The second indication was GBS screening in pregnant women. It represented 166 (16.2%) of analysed samples, which represented 74% of pregnant women included in the study. Finally, STI screening was requested only in 6% of participants.

Because recruitment was conducted directly by health care practitioners, it was not surprising to find good adhesion to clinician examination among the surveyed women. In fact, only a minority (27%) considered the consultation an obstacle to seeking care. However self-collection was well accepted and 84% of participants would recommend the use of VSS. Moreover, 56% of surveyed women stated that VCS substitution by VSS would ensure better follow-up. Only 26% of participants preferred VCS to VSS, mostly because they were not confident with the self-collection process (91%). A recent longitudinal study showed that VSS acceptability improved over time as women became more familiar with the test. We can therefore assume that the proportion of women who prefer VSS will increase. As previously reported, acceptability of VSS was satisfactory among sexually active women surveyed in this study ^43–47^.

### Strenghts and limitations

Conversely to most studies including small subsets of participants, our study benefited from a large cohort (more than a 1000) of women, hence providing an accurate assessment of the added value of VSS for future clinical standard-of-care. This large cohort provided robust assessment of diagnostic performance. In addition, all self-collection results were paired with vaginal classical co-testing. Both samples were collected with the same procedure, treated with the same analytical methodology and within the same delay.

Our recruitment approach excluded infrequently screened women, although previous studies have already reported a similar VSS efficacy and acceptability among within that population ^8,17^. The novelty of our study focuses on the utility of VSS to improve gynaecologic monitoring regarding the challenge of pelvic examination.

The study could have been strengthened by assessing VSS adequacy (ie β-globing testing), although high agreements between VCS and VSS comforted its validity.

Another limitation of our study is that α-risk was not corrected for multiplicity. However, this limitation should be relativized according to the design of the study (non-inferiority of VSS vs. VCS), the fact that observed absolute prevalence’s of VSS are higher than VCS il all cases (except for BV), and that all p-value are less than 0.05 (among which only few are between 0.01 and 0.05)^48,49^

### Implication for practice

Since 2014, in the United States, the NIH and the Centers for Disease Control and Prevention (CDC) recommend the use of VSS for screening of CT and NG by nucleic acid amplification techniques ^8,50^. In addition, they encourage the research community to initiate clinical trials and obtain evidence needed to use VSS for GI screening in a wide variety of settings. Given its efficacity and acceptability, VSS may be considered as an effective alternative to alleviate gynaecological consultation among women needing vaginal sampling for clinical reasons, antenatal screening, and sexual health examinations. Moreover, VSS may enhance acceptability among under-screened low-income women and thereby improve detection and treatment of GIs and STIs. Another interesting application of VSS would be the reliability to repeat a positive test to ensure clearance of the infection following treatment.

## Conclusion

To date, this is the first large-scale cross-sectional study conducted to evaluate the efficacy of VSS, with more than 1000 participants in a global clinical setting, which also included pregnant women.

This study remains the most exhaustive in screening bacterial and yeast infections, multiple STI agents and asymptomatic GBS carriage. Throughout constant efforts to improve medical care, VSS seems to be a viable alternative to the classic physician sampling. It may be a good option to enhance gynaecological monitoring while also alleviating the need for patient intimacy. This study provides evidence that VSS can be used as a universal specimen for detection of lower genital tract infections in women ^8^.

## Data Availability

Data will be available to researchers who provide a methodologically sound proposal. Requests should be addressed by email to

## CONTRIBUTOR AND GUARANTOR INFORMATION

P Halfon and C Camus conceived, designed, and managed the study; P Halfon, G Pénaranda, and L Molet analysed results of the study and wrote the paper; G Penaranda performed statistical analysis; L Chiche assisted in design and analysis of the study; H Khiri, S Camiade and M Lebsir supervised the laboratory specimen analysis; A Plauzolles revised the English manuscript; E Quarello and B Blanc were the main investigators. All authors have read and approved the final version of the manuscript.

## ACKNOWLEDGMENTS

We would like to thank the following gynaecologists of the Marseille European Hôpital Européen Marseille for their contributions: Pr Bernard Blanc, Dr Michel Conte, Dr Olivia Mouremble, Dr Agnes Giocanti, Dr Laurence Villaret, Dr Marie-Jeanne Ducassou, Dr Olivier Le Touze, Dr Alexandre Lazard, Dr Jean Quilichini, Dr Yves-Jean Bernard, Dr Brice Gurriet and Dr Lison Stefani. We also would like to thank the following practitioners: Dr Quarello and his team of midwives at Hôpital St Joseph, Pôle Parents-enfants Ste Monique (N. Caraplis, M. Perrin, F. Billon, L. Villecroze, N. Senatore, V. Rajaoba and A. Deragopian), midwives from the liberal medical office (E. Lambert and A. Rochette) and gynaecologists from the liberal medical office: Dr Anne Suau-Falabregues, Dr Corinne Hassan, Dr Frederic Thoreau and Dr Bernadette Guiomar Mege. The authors also acknowledge all the clinical research associates, the medical biologists, nurses and technical collaborators from European Alphabio Laboratory and European Hospital of Marseille for their useful collaboration in patient recruitment and sample analysis.

## REFERENCES

1. Taylor BD, Darville T, Haggerty CL. Does Bacterial Vaginosis Cause Pelvic Inflammatory Disease?: Sex Transm Dis. 2013 Feb;40(2):117–22.

2. van Schalkwyk J, Yudin MH, Yudin MH, Allen V, Bouchard C, Boucher M, et al. Vulvovaginitis: Screening for and Management of Trichomoniasis, Vulvovaginal Candidiasis, and Bacterial Vaginosis. J Obstet Gynaecol Can. 2015 Mar;37(3):266– 74.

3. Cazanave C, Manhart LE, Bébéar C. Mycoplasma genitalium, an emerging sexually transmitted pathogen. Med Mal Infect. 2012 Sep;42(9):381–92.

4. Lockhart A, Psioda M, Ting J, Campbell S, Mugo N, Kwatampora J, et al. Prospective Evaluation Of Cervico-Vaginal Self And Cervical Physician-Collection For The Detection Of Chlamydia Trachomatis, Neisseria Gonorrhoeae, Trichomonas Vaginalis, And Mycoplasma Genitalium Infections: Sex Transm Dis. 2018 Jan;1.

5. Schrag SJ, Verani JR. Intrapartum antibiotic prophylaxis for the prevention of perinatal group B streptococcal disease: Experience in the United States and implications for a potential group B streptococcal vaccine. Vaccine. 2013 Aug;31:D20–6.

6. Lawn JE, Bianchi-Jassir F, Russell NJ, Kohli-Lynch M, Tann CJ, Hall J, et al. Group B Streptococcal Disease Worldwide for Pregnant Women, Stillbirths, and Children: Why, What, and How to Undertake Estimates? Clin Infect Dis. 2017 Nov 6;65(suppl_2):S89–99.

7. Graseck AS, Shih SL, Peipert JF. Home versus clinic-based specimen collection for Chlamydia trachomatis and Neisseria gonorrhoeae. Expert Rev Anti Infect Ther. 2011 Feb;9(2):183–94.

8. Hobbs MM, van der Pol B, Totten P, Gaydos CA, Wald A, Warren T, et al. From the NIH: Proceedings of a Workshop on the Importance of Self-Obtained Vaginal Specimens for Detection of Sexually Transmitted Infections: Sex Transm Dis. 2008 Jan;35(1):8–13.

9. Shih SL, Graseck AS, Secura GM, Peipert JF. Screening for sexually transmitted infections at home or in the clinic? Curr Opin Infect Dis. 2011 Feb;24(1):78–84.

10. Fajardo-Bernal L, Aponte-Gonzalez J, Vigil P, Angel-Müller E, Rincon C, Gaitán HG, et al. Home-based versus clinic-based specimen collection in the management of Chlamydia trachomatis and Neisseria gonorrhoeae infections. Cochrane STI Group, editor. Cochrane Database Syst Rev [Internet]. 2015 Sep 29 [cited 2019 Apr 30]; Available from: http://doi.wiley.com/10.1002/14651858.CD011317.pub2

11. Let’s Take A “Selfie”: Self-Collected Samples for STIs. Sex Transm Dis. 2018 Jan;1.

12. Sancho-Garnier H, Tamalet C, Halfon P, Leandri FX, Retraite LL, Djoufelkit K, et al. HPV self-sampling or the Pap-smear: A randomized study among cervical screening nonattenders from lower socioeconomic groups in France: HPV self-sampling or the Pap-smear for screening among nonattenders women? Int J Cancer. 2013 May;n/a-n/a.

13. Ilangovan K, Kobetz E, Koru-Sengul T, Marcus EN, Rodriguez B, Alonzo Y, et al. Acceptability and Feasibility of Human Papilloma Virus Self-Sampling for Cervical Cancer Screening. J Womens Health. 2016 Sep;25(9):944–51.

14. Nelson EJ, Maynard BR, Loux T, Fatla J, Gordon R, Arnold LD. The acceptability of self-sampled screening for HPV DNA: a systematic review and meta-analysis. Sex Transm Infect. 2017 Feb;93(1):56–61.

15. Scarinci IC, Litton AG, Garcés-Palacio IC, Partridge EE, Castle PE. Acceptability and Usability of Self-Collected Sampling for HPV Testing Among African-American Women Living in the Mississippi Delta. Womens Health Issues. 2013 Mar;23(2):e123–30.

16. Montealegre JR, Mullen PD, L. Jibaja-Weiss M, Vargas Mendez MM, Scheurer ME. Feasibility of Cervical Cancer Screening Utilizing Self-sample Human Papillomavirus Testing Among Mexican Immigrant Women in Harris County, Texas: A Pilot Study. J Immigr Minor Health. 2015 Jun;17(3):704–12.

17. Lunny C, Taylor D, Hoang L, Wong T, Gilbert M, Lester R, et al. Self-Collected versus Clinician-Collected Sampling for Chlamydia and Gonorrhea Screening: A Systemic Review and Meta-Analysis. Greub G, editor. PLOS ONE. 2015 Jul 13;10(7):e0132776.

18. Blackwelder WC. “Proving the null hypothesis” in clinical trials. Control Clin Trials. 1982 Dec;3(4):345–53.

19. Schwebke JR, Hillier SL, Sobel JD, McGregor JA, Sweet RL. Validity of the vaginal gram stain for the diagnosis of bacterial vaginosis. Obstet Gynecol. 1996 Oct;88(4 Pt 1):573–6.

20. Landis JR, Koch GG. The Measurement of Observer Agreement for Categorical Data. Biometrics. 1977 Mar;33(1):159.

21. Schachter J, Chernesky MA, Willis DE, Fine PM, Martin DH, Fuller D, et al. Vaginal swabs are the specimens of choice when screening for Chlamydia trachomatis and Neisseria gonorrhoeae: results from a multicenter evaluation of the APTIMA assays for both infections. Sex Transm Dis. 2005 Dec;32(12):725–8.

22. Garrow SC. The diagnosis of chlamydia, gonorrhoea, and trichomonas infections by self obtained low vaginal swabs, in remote northern Australian clinical practice. Sex Transm Infect. 2002 Aug 1;78(4):278–81.

23. Wroblewski JKH, Manhart LE, Dickey KA, Hudspeth MK, Totten PA. Comparison of transcription-mediated amplification and PCR assay results for various genital specimen types for detection of Mycoplasma genitalium. J Clin Microbiol. 2006 Sep;44(9):3306–12.

24. Rompalo AM, Gaydos CA, Shah N, Tennant M, Crotchfelt KA, Madico G, et al. Evaluation of use of a single intravaginal swab to detect multiple sexually transmitted infections in active-duty military women. Clin Infect Dis Off Publ Infect Dis Soc Am. 2001 Nov 1;33(9):1455–61.

25. Smith KS, Tabrizi SN, Fethers KA, Knox JB, Pearce C, Garland SM. Comparison of conventional testing to polymerase chain reaction in detection of Trichomonas vaginalis in indigenous women living in remote areas. Int J STD AIDS. 2005 Dec;16(12):811–5.

26. Des Marais AC, Zhao Y, Hobbs MM, Sivaraman V, Barclay L, Brewer NT, et al. Home Self-Collection by Mail to Test for Human Papillomavirus and Sexually Transmitted Infections: Obstet Gynecol. 2018 Nov;1.

27. Tronstein E. Genital Shedding of Herpes Simplex Virus Among Symptomatic and Asymptomatic Persons With HSV-2 Infection. JAMA. 2011 Apr 13;305(14):1441.

28. Boskey ER, Atherly-Trim SA, O’Campo PJ, Strobino DM, Misra DP, Misra P. Acceptability of a self-sampling technique to collect vaginal smears for gram stain diagnosis of bacterial vaginosis. Womens Health Issues Off Publ Jacobs Inst Womens Health. 2004 Feb;14(1):14–8.

29. Yen S, Shafer M-A, Moncada J, Campbell CJ, Flinn SD, Boyer CB. Bacterial vaginosis in sexually experienced and non-sexually experienced young women entering the military. Obstet Gynecol. 2003 Nov;102(5 Pt 1):927–33.

30. Bradshaw CS, Morton AN, Hocking J, Garland SM, Morris MB, Moss LM, et al. High recurrence rates of bacterial vaginosis over the course of 12 months after oral metronidazole therapy and factors associated with recurrence. J Infect Dis. 2006 Jun 1;193(11):1478–86.

31. Strauss RA, Eucker B, Savitz DA, Thorp JM. Diagnosis of bacterial vaginosis from self-obtained vaginal swabs. Infect Dis Obstet Gynecol. 2005 Mar;13(1):31–5.

32. Khan Z, Bhargava A, Mittal P, Bharti R, Puri P, Khunger N, et al. Evaluation of reliability of self-collected vaginal swabs over physician-collected samples for diagnosis of bacterial vaginosis, candidiasis and trichomoniasis, in a resource-limited setting: a cross-sectional study in India. BMJ Open. 2019 Aug;9(8):e025013.

33. Barnes P, Vieira R, Harwood J, Chauhan M. Self-taken vaginal swabs versus clinician-taken for detection of candida and bacterial vaginosis: a case-control study in primary care. Br J Gen Pract. 2017 Dec;67(665):e824–9.

34. Kashyap B, Singh R, Bhalla P, Arora R, Aggarwal A. Reliability of self-collected versus provider-collected vaginal swabs for the diagnosis of bacterial vaginosis. Int J STD AIDS. 2008 Aug;19(8):510–3.

35. van de Wijgert J, Altini L, Jones H, de Kock A, Young T, Williamson A-L, et al. Two Methods of Self-Sampling Compared to Clinician Sampling to Detect Reproductive Tract Infections in Gugulethu, South Africa: Sex Transm Dis. 2006 Aug;33(8):516– 23.

36. Vergers-Spooren HC, van der Meijden WI, Luijendijk A, Donders G. Self-Sampling in the Diagnosis of Recurrent Vulvovaginal Candidosis: J Low Genit Tract Dis. 2013 Apr;17(2):187–92.

37. Mercer BM, Taylor MC, Fricke JL, Baselski VS, Sibai BM. The accuracy and patient preference for self-collected group B Streptococcus cultures. Am J Obstet Gynecol. 1995 Oct;173(4):1325–8.

38. Hicks P, Diaz-Perez MJ. Patient Self-Collection of Group B Streptococcal Specimens During Pregnancy. J Am Board Fam Med. 2009 Mar 1;22(2):136–40.

39. Arya A, Cryan B, O’Sullivan K, Greene RA, Higgins JR. Self-collected versus health professional-collected genital swabs to identify the prevalence of group B streptococcus: A comparison of patient preference and efficacy. Eur J Obstet Gynecol Reprod Biol. 2008 Jul;139(1):43–5.

40. Forney LJ, Gajer P, Williams CJ, Schneider GM, Koenig SSK, McCulle SL, et al. Comparison of self-collected and physician-collected vaginal swabs for microbiome analysis. J Clin Microbiol. 2010 May;48(5):1741–8.

41. Price D, Shaw E, Howard M, Zazulak J, Waters H, Kaczorowski J. Self-Sampling for Group B Streptococcus in Women 35 to 37 Weeks Pregnant Is Accurate and Acceptable: A Randomized Cross-Over Trial. J Obstet Gynaecol Can. 2006 Dec;28(12):1083–8.

42. Ness RB, Kip KE, Hillier SL, Soper DE, Stamm CA, Sweet RL, et al. A Cluster Analysis of Bacterial Vaginosis–associated Microflora and Pelvic Inflammatory Disease. Am J Epidemiol. 2005 Sep 15;162(6):585–90.

43. Chernesky M, Jang D, Gilchrist J, Randazzo J, Elit L, Lytwyn A, et al. Ease and Comfort of Cervical and Vaginal Sampling for Chlamydia trachomatis and Trichomonas vaginalis with a New Aptima Specimen Collection and Transportation Kit. J Clin Microbiol. 2014 Feb 1;52(2):668–70.

44. Paudyal P, Llewellyn C, Lau J, Mahmud M, Smith H. Obtaining Self-Samples to Diagnose Curable Sexually Transmitted Infections: A Systematic Review of Patients’ Experiences. Clark JL, editor. PLOS ONE. 2015 Apr 24;10(4):e0124310.

45. Doshi JS, Power J, Allen E. Acceptability of chlamydia screening using self-taken vaginal swabs. Int J STD AIDS. 2008 Aug;19(8):507–9.

46. Fielder RL, Carey KB, Carey MP. Acceptability of sexually transmitted infection testing using self-collected vaginal swabs among college women. J Am Coll Health J ACH. 2013;61(1):46–53.

47. Odesanmi TY, Wasti SP, Odesanmi OS, Adegbola O, Oguntuase OO, Mahmood S. Comparative effectiveness and acceptability of home-based and clinic-based sampling methods for sexually transmissible infections screening in females aged 14-50 years: a systematic review and meta-analysis. Sex Health. 2013 Dec;10(6):559–69.

48. Althouse AD. Adjust for Multiple Comparisons? It’s Not That Simple. Ann Thorac Surg. 2016 May;101(5):1644–5.

49. Rothman KJ. No adjustments are needed for multiple comparisons. Epidemiol Camb Mass. 1990 Jan;1(1):43–6.

50. Papp JR, Schachter J, Gaydos CA, Van B. Recommendations for the Laboratory-Based Detection of Chlamydia trachomatis and Neisseria gonorrhoeae — 2014. 2014;34.

