## Supplemental File 1 for "Acceptability and efficacy of vaginal self-sampling for genital infection and bacterial vaginosis: A large, cross-sectional, non-inferiority trial"

**Supplementary File 1**

### APV Protocol

### Questionnaire about vaginal self-sampling

**1-Which type of sampling did you preferred?**

- □ Vaginal Self-Sampling, why? (thick one or more boxes)
  - □ Easy and less embarrassing
  - □ Is not opposed to my cultural norms
  - □ Facilitate monitoring of genital infections
  - □ Financial reason (free of charge)
  - □ Other:
- □ Vaginal Classic-Sampling, why? (thick one or more boxes)
  - □ The practitioner has more experience than I do, and the sampling may be of better quality
  - □ Feel not comfortable with vaginal self-sampling
  - □ Vaginal self-sampling was more unpleasant or painful than classic sampling
  - □ It was hard to perform vaginal self-sampling
- □ Both

**2-Accordinf to you, is a medical appointment to perform vaginal sampling is an obstacle to gynecological monitoring? (thick only one box)**

- □ Yes; do you prefer that your vaginal sampling is performed by: (thick one or more boxes)
  - □ Medical biologist at the medical laboratory?
  - □ Yourself at the medical laboratory?
  - □ Yourself at home?
- □ No

**3-Would you recommend vaginal self-sampling?**

- □ Yes
- □ No

**4-In the case where vaginal self-sampling replaces classical sampling, would it encourage you to be monitored more regularly?**

□ Yes

□ No

**5-What is your highest level of education?**

□ None (elementary school)

□ Basic (secondary school level)

□ Medium (high school level)

□ High (university level)
